# Emergence of *mcr-9.1* in ESBL-producing Clinical Enterobacteriaceae in Pretoria, South Africa: Global Evolutionary Phylogenomics, Resistome and Mobilome

**DOI:** 10.1101/2019.12.24.19015784

**Authors:** John Osei Sekyere, Nontuthuko E. Maningi, Lesedi Modipane, Nontombi Marylucy Mbelle

**Author notes:** **Direct correspondence** to John Osei Sekyere, PhD. Department of Medical Microbiology, University of Pretoria.

## Abstract

**Background:** Extended-spectrum β-lactamase (ESBL)-producing *Enterobacteriaceae* are critical-priority pathogens that cause substantial fatalities. With the emergence of mobile *mcr* genes mediating resistance to colistin in Enterobacteriaceae, clinicians are now left with little therapeutic options.

**Methods:** Eleven clinical Enterobacteriaceae strains with resistance to cephems and/or colistin were genomically analysed to determine their resistome, mobilome, and evolutionary relationship to global strains. The global phylogenomics of *mcr-9.1-*bearing genomes were further analysed.

**Results & conclusion:** Ten isolates were ESBL positive. The isolates were multidrug-resistant and phylogenetically related to global clones, but distant from local strains. Multiple resistance genes, including *bla*_CTX-M-15_ *bla*_TEM-1_ and *mcr-9.1* were found in single isolates; IS*Ec9*, IS*19*, and Tn*3* transposons bracketed *bla*_CTX-M-15_ and *bla*_TEM-1_. Common plasmid types included IncF, IncH and ColRNAI. Genomes bearing *mcr-9.1* clustered into six main phyletic groups (A-F), with those of this study belonging to clade B. *Enterobacter sp*. and *Salmonella sp*. are the main hosts of *mcr-9.1* globally, albeit diverse promiscuous plasmids disseminate *mcr-9.1* across different bacterial species. Emergence of *mcr-9.1* in ESBL-producing Enterobacteriaceae in South Africa is worrying due to the restricted therapeutic options. Intensive One Health molecular surveillance might discover other *mcr* alleles and inform infection management and antibiotic choices.

## Introduction

Enterobacteriaceae producing extended-spectrum β-lactamases (ESBLs) are categorised as critical priority 1 pathogens requiring urgent attention with regards to the development of new antimicrobials ^1^. Owing to the fatalities caused by ESBL-positive Enterobacteriaceae, carbapenems were introduced as last-resort antibiotics into clinical practice, which unfortunately led to the proliferation of strains expressing carbapenem resistance through ESBLs/AmpCs hyper-expression, porin downregulation and carbapenemase production ^2–4^. Subsequently, colistin was re-introduced into clinical medicine as a last-resort agent to manage infections resistant to carbapenems, other β-lactams and other antibiotics ^5,6^.

Inexorably, the increased use of colistin also resulted in the emergence of colistin resistance ^3,5,7,8^. Since 2016, the discovery of a mobile colistin resistance gene, *mcr-1*, has enhanced the dissemination of colistin resistance worldwide ^5,9,10^. Several alleles of the *mcr-1* gene such as *mcr-1* to *mcr-9*, have been also been discovered, mainly in Europe and China in several Enterobacteriaceae species including *Escherichia coli, Enterobacter spp. Klebsiella spp*., *Salmonella spp*., etc. ^11–14^.

Worryingly, increasing reports of isolates co-harbouring carbapenemases, ESBLs and *mcr* genes are being reported worldwide and on self-transmissible plasmids ^4,15–19^. Such strains could be pandrug resistant, restricting therapeutic options for clinicians and threatening public health ^6,8^.

In this report, we describe for the first time the emergence of *mcr-9.1* colistin resistance gene in South Africa and Africa as well as the co-occurrence of *mcr-9.1* with ESBLs in *Enterobacter hormaechei*. We further undertake a global phylogenomic analyses of *Citrobacter freundii, Enterobacter hormaechei, Klebsiella variicola*, and *Providencia/Proteus spp*., alongside a global evolutionary analysis of *mcr-9.1*. These findings call for advanced One Health genomic surveillance to contain such multi-drug resistant strains from further dissemination.

## Methods

### Clinical bacterial specimens and antibiotic sensitivity testing

An old collection of clinical specimens (n=72) that were obtained in 2013 at a referral laboratory from patients having drug-resistant infections at an academic teaching hospital were analysed ^20–22^. These patients suffered from several different diseases and the specimens were obtained from different body sites. The specimens were cultured on blood agar overnight and pure colonies were transferred to freshly prepared Mueller-Hinton plates for another 24-hour incubation. The fresh cultures were then used for antimicrobial sensitivity testing and species identification on a MiscroScan Walkaway system. The isolates were further tested for ESBL production using the double-disk synergy testing ^23^.

Enterobacteriaceae species having reduced sensitivity to cephems and colistin were further selected for genomic analyses.

### Whole-genome sequencing

Eleven isolates belonging to Enterobacteriaceae species that are rarely isolated from the referral laboratory were cultured for 24 hours. Their genomic DNA were subsequently isolated using the ZR Fungal/Bacterial DNA Mini-Prep kit (Zymo Research, USA). Briefly, 200bp libraries were prepared from the gDNA and size-selected using 2% agarose gel and Pippen prep (Sage Science, Beverly, MA, USA). The libraries were barcoded and pooled together for sequencing on an Ion Proton (ThermoFisher, Waltham, MA, USA). SPAdes assembler was used to assemble the generated raw reads de novo.

### Bioinformatic and phylogenomic analyses

The assembled sequences were deposited at Genbank and annotated with PGAP ^24^. The resistome of the isolates were determined from the Isolates Browser database of NCBI (https://www.ncbi.nlm.nih.gov/pathogens/isolates#/search/) whilst the mobilome were manually curated from the gff files from Genbank. The isolates’ multi-locus sequence types (MLST) sequence types were identified with the MLST database at CGE (http://cge.cbs.dtu.dk/services/MLST/). INTEGRALL (http://cge.cbs.dtu.dk/services/MLST/). INTEGRALL (http://integrall.bio.ua.pt/) was used to identify the integrons and gene cassettes within the genomes whilst PlasmidFinder (http://cge.cbs.dtu.dk/services/PlasmidFinder/) was used to identify the plasmid replicons. Contigs bearing the *mcr9.1* genes were BLASTed (using nucleotide BLAST) to identify genomes with closest nucleotide identity, which were used to draw distance trees using the Fast Minimum Evolution method at a Maximum Sequence Difference of 0.75. The genomes of *Citrobacter freundii, Enterobacter hormaechei, Klebsiella variicola, Providencia spp*. and *Proteus mirabilis* were curated from PATRIC (https://www.patricbrc.org) and filtered to remove poor genomes. The filtered genomes were then run through RAxML to draw maximum-likelihood trees that were annotated using Figtree (http://tree.bio.ed.ac.uk/software/figtree/).

## Results

### Demographics, genome characteristics & resistance phenome

The specimens were mainly sourced from six males and four females, aged between 18 and 61, and hospitalized at a single tertiary academic hospital in Pretoria, South Africa (Figure 1; Table 1). Except for the carbapenems, cephalothin, and combinations of clavulanate and tazobactam with cefotaxime/ceftazidime and piperacillin respectively, most of the isolates were resistant to all the β-lactams. Moreover, reduced resistance to piperacillin-tazobactam (in EC009, EC010, and K001) and sensitivity to aztreonam and cefepime (CF004), and cefoxitin (EC015, K001, K006, K130, K063 and PM005) were observed. All the isolates were phenotypically positive for ESBL production, except CF004 (Supplementary Table S1 and Table 1). None of the isolates was resistant to amikacin. CF004 was susceptible to all the non-β-lactam antibiotics whilst the remaining strains were resistant to almost all the non-β-lactam antibiotics except tigecycline and Fosfomycin. Four of the isolates were resistant to colistin (Supplementary Table S1).

**Table 1.**
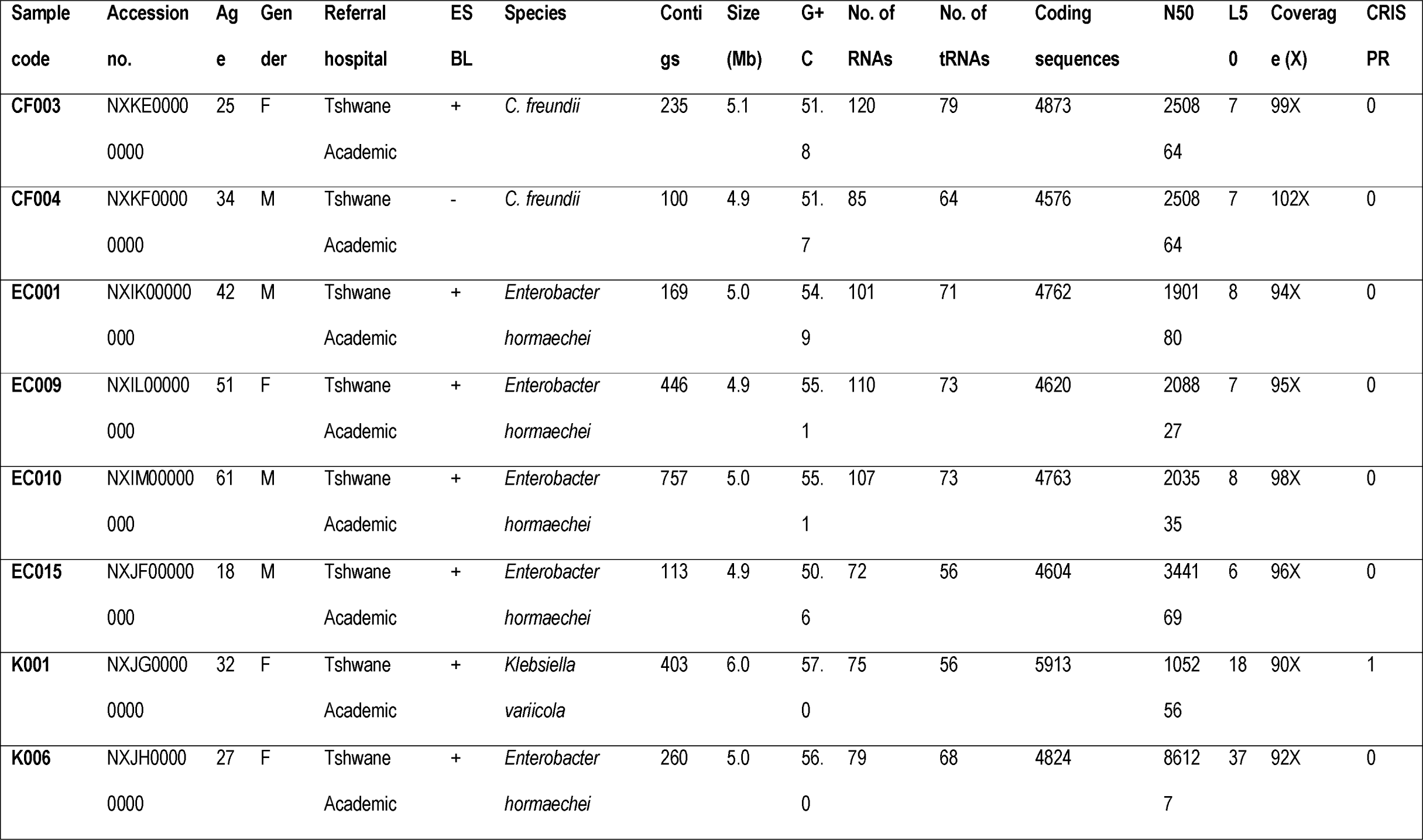
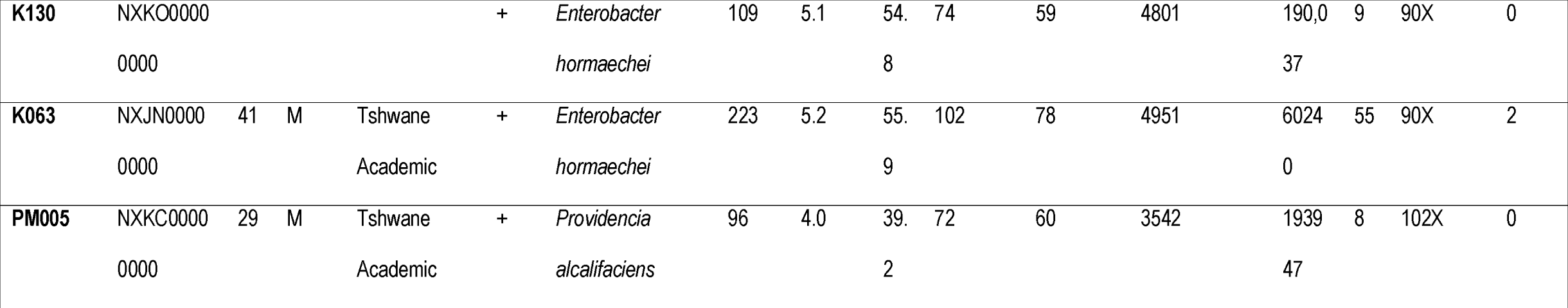
Demographic and genomic characteristics of the isolates.

**Figure 1.**
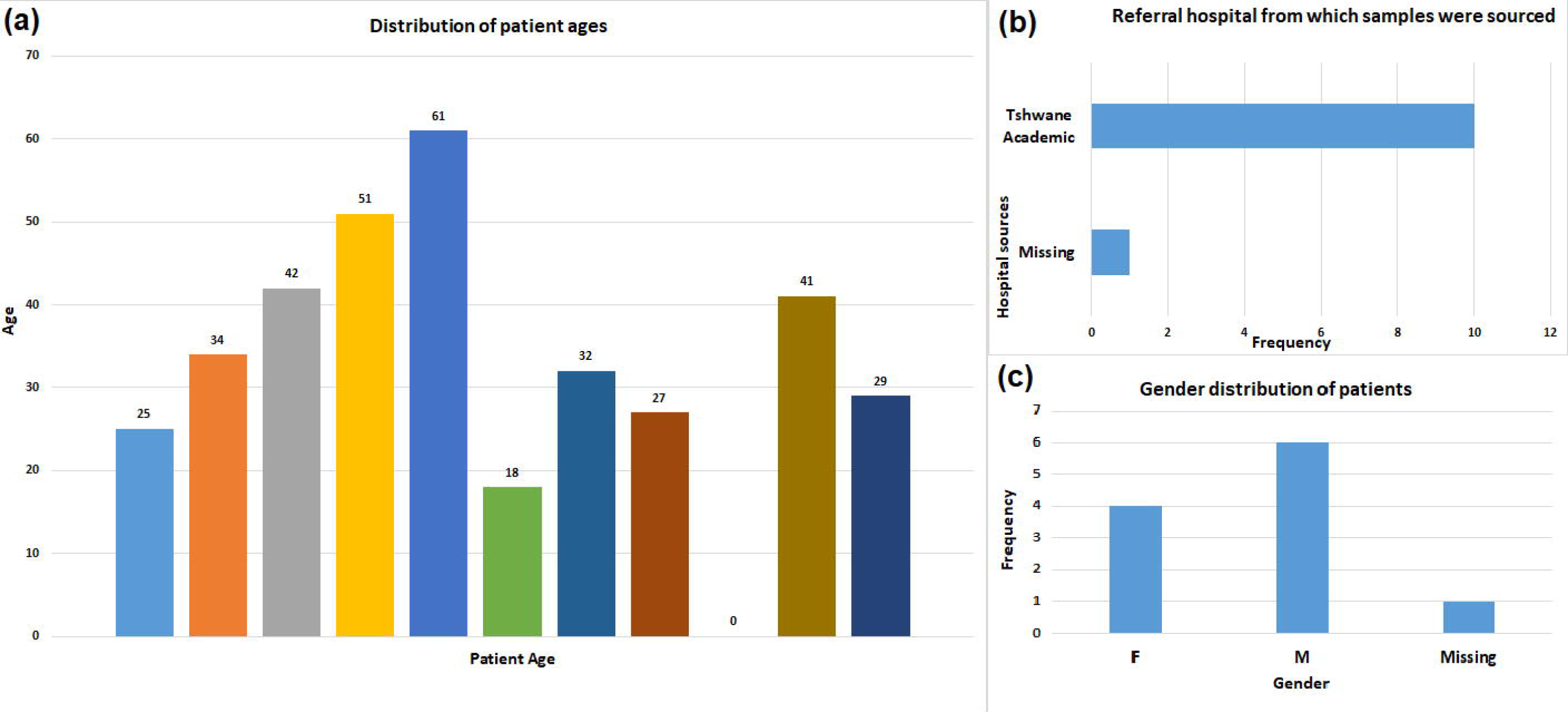
Demographic characteristics of patients from whom the specimens were sourced from. The patients’ ages (A), the hospital where they were hospitalized (B) and their gender (C) are shown herein. These were calculated directly from the source documents populated in Microsoft Excel.

The draft genome sizes of the genomic sequences ranged from 4Mb to 6Mb, with PM005 having the lowest GC content (39.2) and coding sequences (3542). The genome coverage ranged from 90X to 102X whilst CRISPR arrays were identified in only two isolates (K001 and K063) (Table 1; Supplementary dataset 1).

The isolates were all multi-drug resistant and agreements as well as discrepancies between the resistance phenotypes (phenomes) and resistance genes (resistomes) were observed (Table S1). In particular, CF003 and CF004 had only *bla*_CMY_ and *aac(6’)-If* or *fosA7* genes, but expressed multidrug resistance to several antibiotics for which no resistance gene were identified. Such observations were also made with the presence of *fosA* genes in the isolates and the absence of fosfomycin resistance in almost all the strains as well as the presence of chromosomal AmpCs such as ACT, LEN and CMY, in EC015, K001, K006, K130, K063 and PM005, which were all susceptible to cefoxitin (Supplementary Table S1). Notable was the absence of colistin resistance in K006 and K130, which had the *mcr-9.1* gene; comparative genomic analyses showed that the colistin-susceptible and -resistant strains shared the same chromosomal mutations in *mgrB* and *pmrB* (Table 2). The absence of carbapenem resistance was corroborated by the absence of any carbapenemase gene whilst the ESBL-positive phenotypes agreed with the ESBL genes identified in the genomes (Supplementary Table S1 and Figure 2).

**Table 2.**
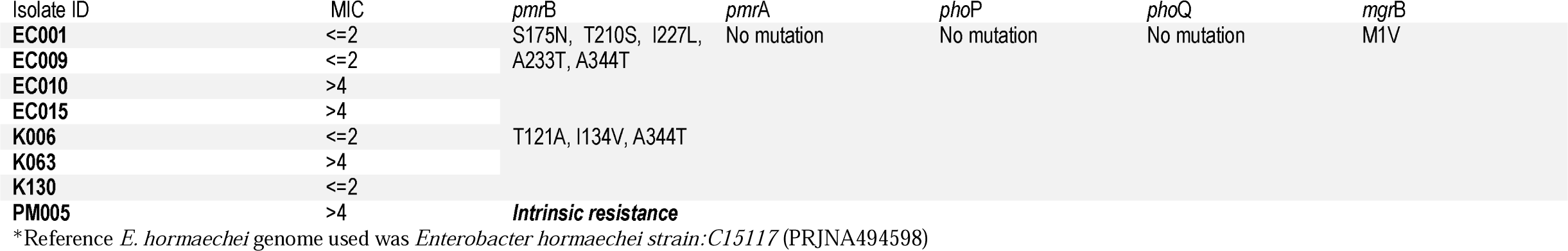
Point mutations on the colistin chromosomal resistance genes of the E. hormaechei isolates from South Africa

**Figure 2.**
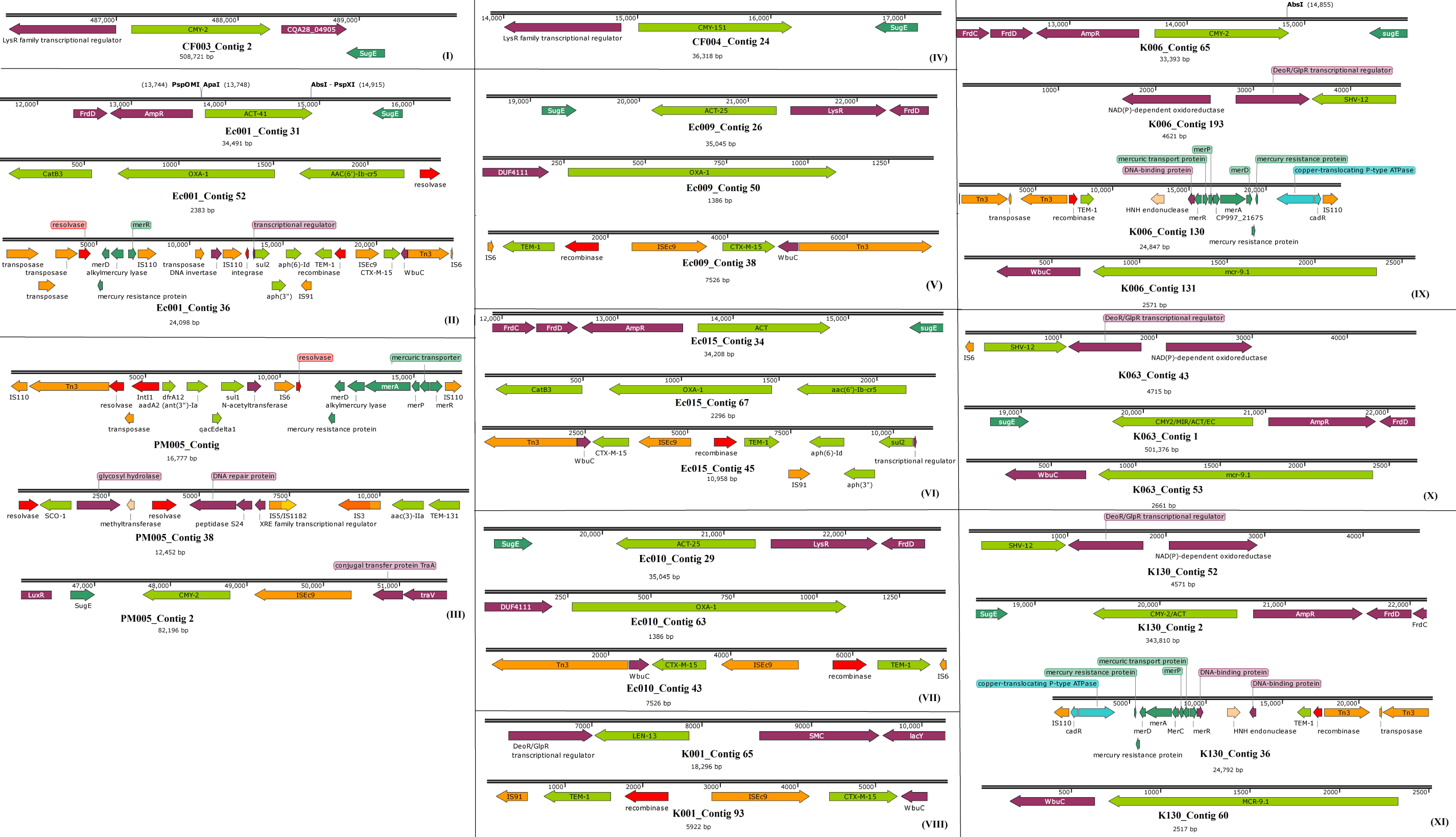
Genetic environment of β-lactamase and *mcr-9.1* genes and evolutionary epidemiology of *mcr-9.1*. The mobile genetic elements viz., integrons/recombinases (red arrows) and transposons/ISs (orange arrows) bracketing the resistance genes (green arrows) are shown for the various isolates under panels I to XI in 2**A**. The evolutionary distance and relationship between the contigs bearing the *mcr-9.1* and highly similar genomes obtained from Genbank are shown in three different trees labelled A (K006), B (K130) and C (K063) in 2**B**. The contigs from this study are labelled red and the genomes of closest alignment are labelled either green (for A|K006 and B|K130) or blue (for C|K063). The various phyletic groupings that form a separate clade are labelled from A to F under different colours to show the evolutionary trajectory of *mcr-9.1*.

### Resistome, mobilome and evolutionary epidemiology of mcr-9

CF003 and CF004 had the least number of resistance genes, with no ESBL genes. Dominant ESBLs in the other nine isolates included CTX-M-15, TEM-1, OXA-1, OXA-9 and ACT (Supplementary Table S1; Supplementary dataset 1). The genetic environment of the contigs on which the AmpC genes such as *bla*_CMY_, *bla*_ACT_, *bla*_LEN_ and *bla*_LAP_ were located strongly suggested their presence on chromosomes. Notably, the *bla*_CMY_ in PM005 was in close synteny with *ISEc9* (Fig. 2AIII). *bla*_CTX-M-15_ and *bla*_TEM-1_ ESBLs were almost always found on the same contig, respectively bracketed by IS*Ec9* and a Tn3 transposon and IS*19* and an integrase/recombinase. In particular, a*ph(6’)-Id:aph(3’’):sul2* resistance genes were commonly found joined to IS*19*, just after *bla*_TEM-1_ in *mcr-9-*negative *E. hormaechei*. In PM005, K006 and K130, the genetic environment of *bla*_TEM-1_ were different (Fig. 2A). The *bla*_OXA_ gene was commonly found between *catB3* and *aac(6’)-Ib* on a class 1 integron as gene cassettes or directly on the chromosome, similar to what was observed for *bla*_SHV,_ *bla*_LEN_, and *bla*_ACT._(Fig. 2A)

A recent allele of the mobile colistin resistance gene, *mcr-9.1*, was identified in three *E. hormaechei* strains, the first to be discovered from Africa, dating back to 2013. *mcr9.1* genes were all found in the immediate environment of a cupin fold metalloprotein, WbuC, similar to *bla*_CTX-M-15_ gene. *mcr9.1* only occurred once with a *bla*_CTX-M-15_ in K063. Nucleotide blast of the contigs viz., K006 contig 131, K063 contig 53 and K130 contig 60, and subsequent phylogenetics using the Fast-Minimum Evolution method showed that the contigs harbouring the *mcr9.1* gene were most closely aligned to plasmid genomes from different Enterobacteriaceae species, with a few chromosomes such as that of *Enterobacter kobei* strain DSM 13645 chromosome being also closely aligned (Fig. 2B). Sequence alignment showed close similarity between the three *mcr-9*-bearing contigs (Supplemental dataset 2), particularly between the contigs of K006 and K130 (Fig. 2B). Notably, both K006 and K130 aligned with very close nucleotide identity with the same genomes, albeit minor differences existed in their nucleotide sequences (Supplemental dataset 2).

Both K006 and K130 were phylogenetically distant from K063 (Fig. 2B), which is further evinced by the different genomes to which they both most closely aligned with and the distance of those genomes from each other in the distance trees (Fig. 2B-2C). The evolutionary epidemiology of the *mcr-9.1-*bearing plasmids or chromosomes is further shown by Fig. 2B-2C, with different plasmid types and chromosomes mediating the spread of this gene within and between species throughout the world. The various *mcr-9.1-*bearing genomes (plasmids or chromosomes) clustered phylogenetically into six, herein labelled as A-F, with *Enterobacter sp*. 18A13 plasmid pECC18A13-1 DNA from Japan seeming to be the earliest ancestor and *Enterobacter hormaechei* strain S13 plasmid pSHV12-1301491 from the USA seeming to be the most recent. All the *mcr-9.1* contigs in this study fell within the B phylogenetic cluster, although the genomes that aligned most closely with these contigs changed their phylogenetic clustering between K006/K130 and K063 (Fig. 2B-2C).

Several integrons bearing gene cassettes of antibiotic resistance genes were identified in various contigs in the isolates (Table 3), with none being detected in the *C. freundii* strains. PM005 (*Providencia alcalifaciens*) had a unique integron, In27, whilst K001 (*K. variicola*) and four *E. hormaechei* strains harboured In*191*. K006, K063 and K130, originating from different patients and having the *mcr-9.1* gene, harboured the same integrons and very similar gene cassettes. EC009 and Ec010 had the same class 1 integron and gene cassettes and belonged to the same clone (Table 3). Most of the isolates had multiple plasmid replicons, with ColRNAI being the commonest; ColRNAI was also found in the *mcr-9.1* strains. The isolates were further grouped using their pMLST (plasmid multi-locus sequence typing). Except for CF004 and PM005, which had no pMLST but contained an A/C_2_ replicon, IncHI2[ST-1] and IncF subtypes were the main plasmid types. Notably, all the *mcr-9.1* strains had an IncHI2[ST-1] plasmid whilst two, K006 and K063, co-haboured IncF sub-type plasmids. Although the resistance genes within strains having the same plasmid types were different, core resistance genes with same genetic environment were present (Fig. 2A; Table 4).

**Table 3.**
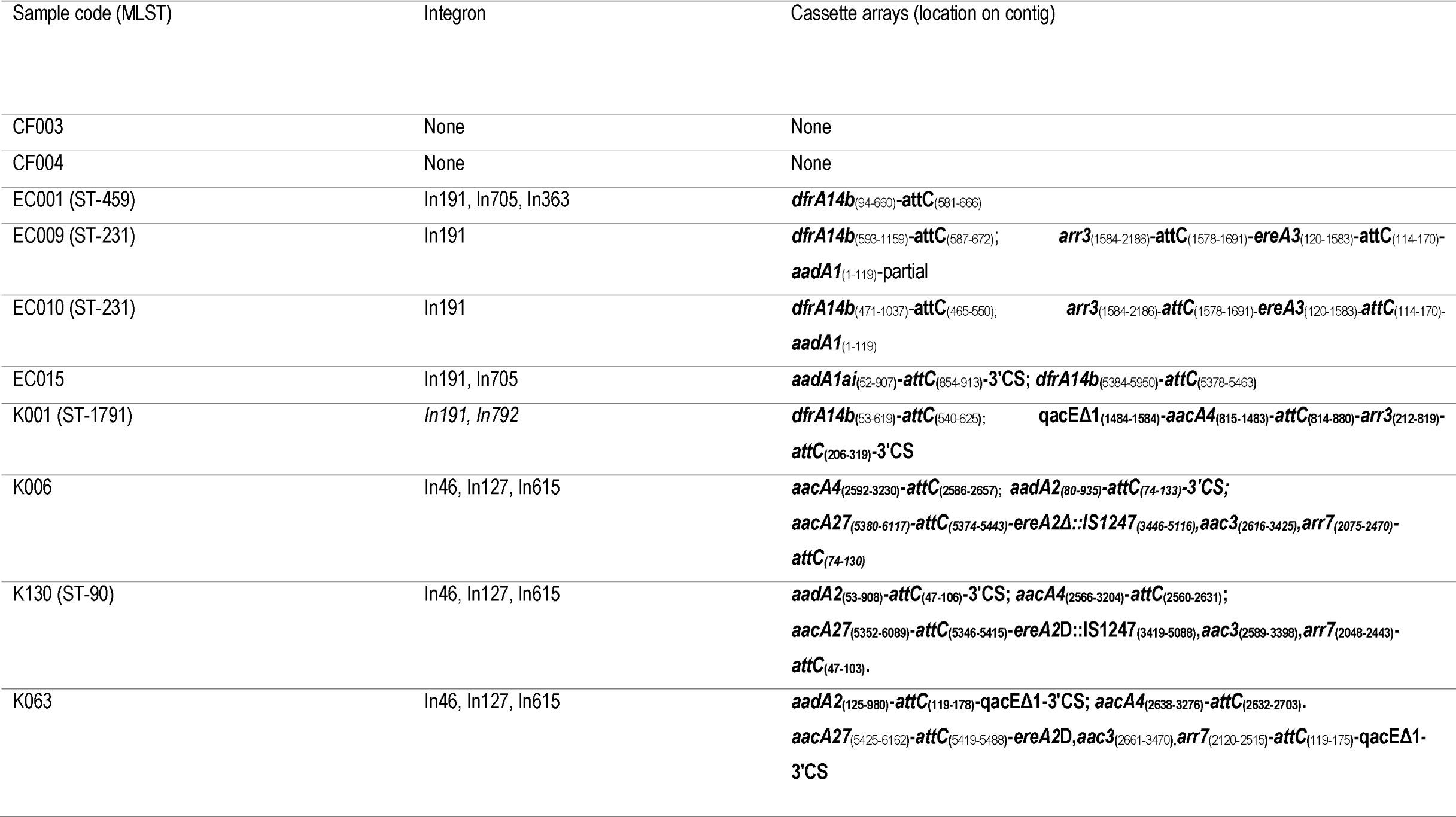
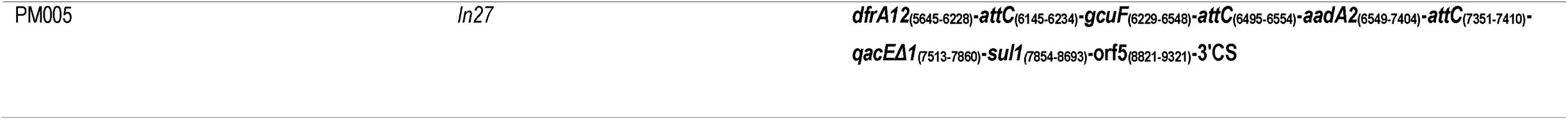
Gene cassettes and integrons found in the isolates

**Table 4.**
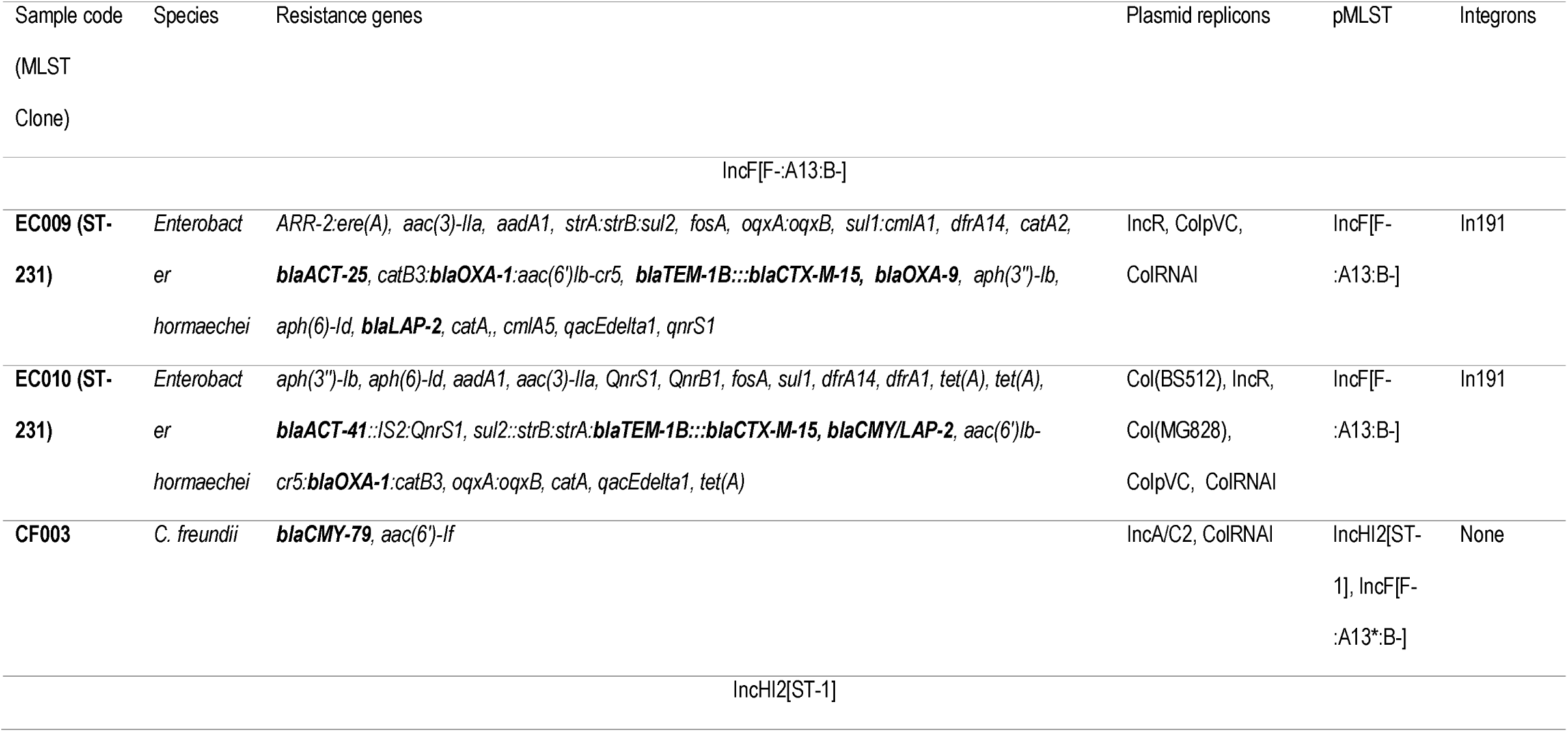
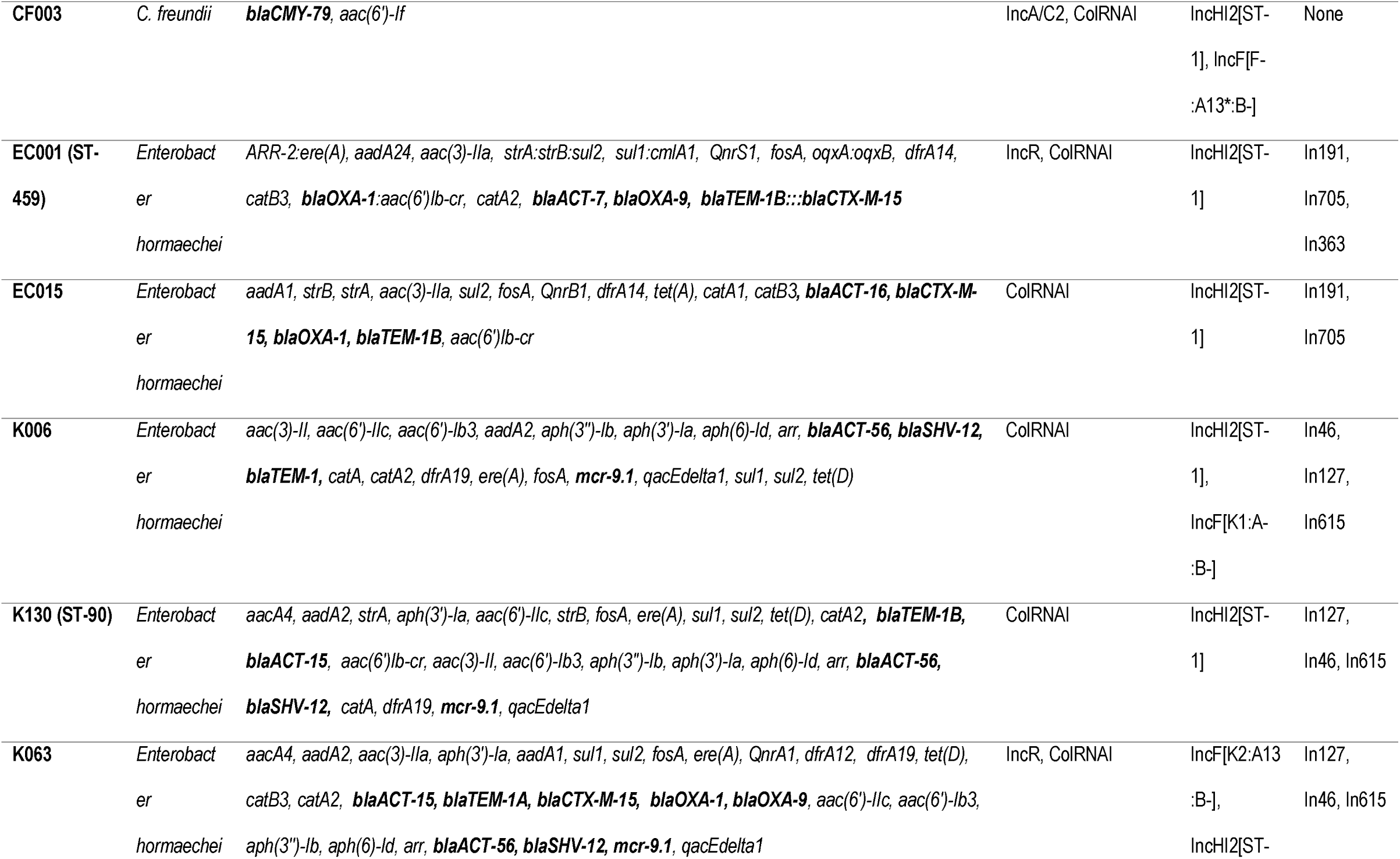
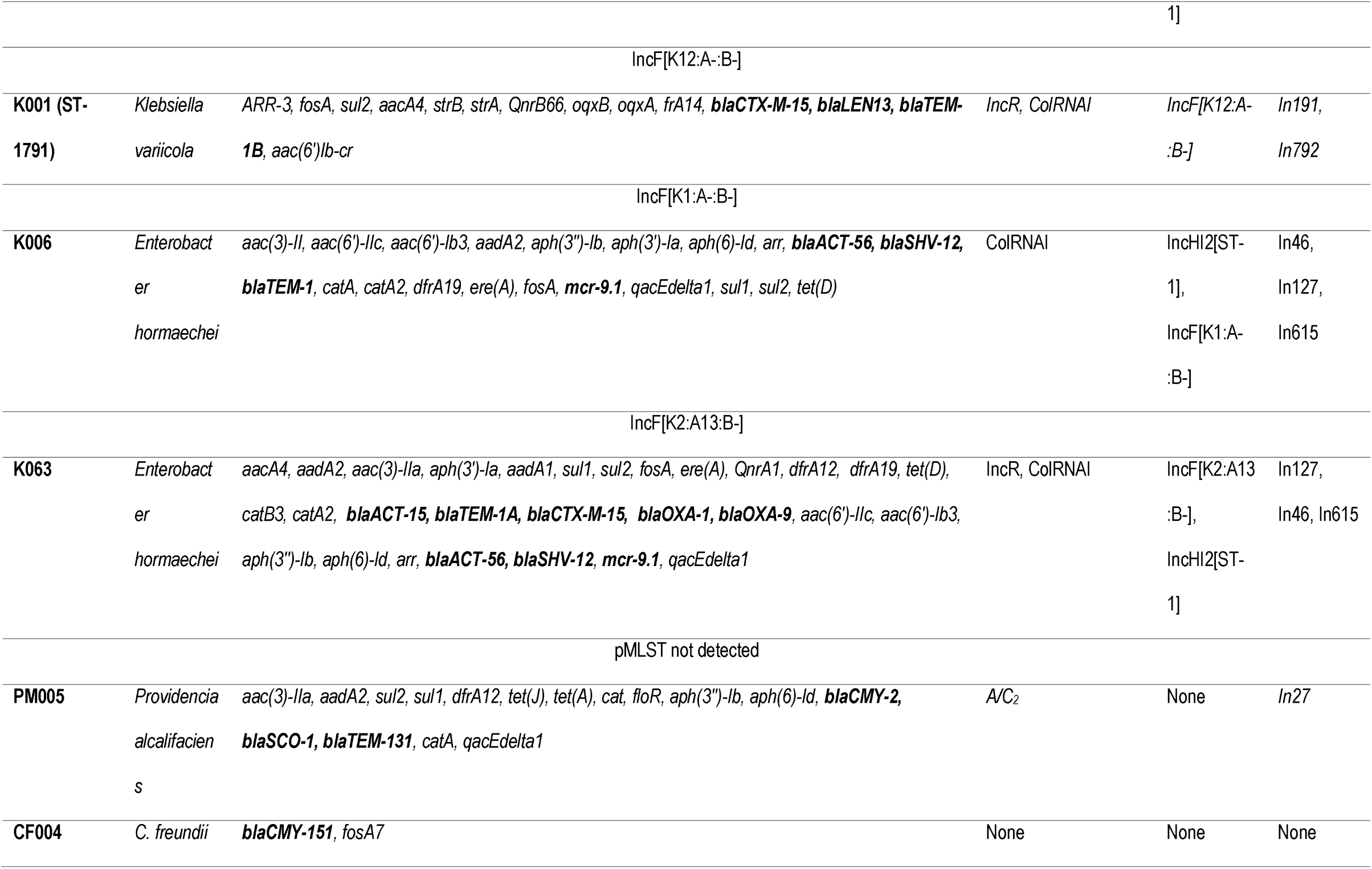
Resistome and mobilome characteristics of the isolates

### Phylogenomics

The *C. freundii* strains were not related to any South African or African strain, but to American and European strains. CF004 and CF003 were not related to each other, but CF004 was of the same clone as ***C. freundii* CF_12_ST_92** [*aac(3)-IIa, aac(6’)-Ib4, aadA1, aph(6)-Id, arr-2*, ***bla***_**CMY-48**_, ***bla***_**OXA**_, ***bla***_**VIM-1**_, *catB2, dfrA14, dfrB1, mph(A), qacE, qnrA1 sul1*] from Spain and of the same clade as strains **705SK3** [***bla***_**CMY-75**_, ***bla***_**OXA-48**_] from Switzerland and **CF_13_ST_93** [*aac(6’)-Ib4, aadA1, aph(3’’)-Ib, aph(6)-Id*, ***bla***_**CMY-75**_, ***bla***_**VIM-1**_, *catB2, dfrA14, dfrB1, qacE*Δ*1, qnrB10, sul1, sul2*] from Spain. CF003 was of the same clade as **MGH142** [***bla***_**CMY**_], **UMH16** [***bla***_**CMY**_], **MGH141** [*aadA2, ant(2’’)-Ia, aph(3’)-II*, ***bla***_**CARB-2**_, ***bla***_**CMY**_, ***bla***_**KPC-3**_, *ble, catB11, catB3, cmlA1, dfrA19, mph(E), msr(E), qacE*Δ*1*] sul1], and **CRE20** [***bla***_**CMY**_]) from USA (Fig. 3).

**Figure 3.**
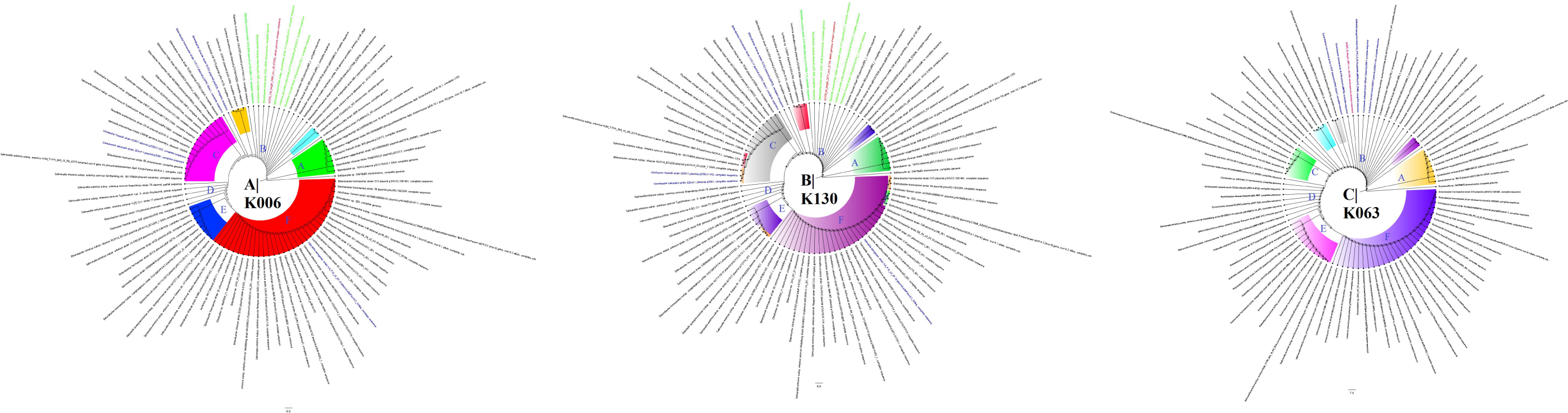
Global phylogenomics of *Citrobacter freundii* strains obtained from PATRIC/GENBANK. The relationship of the two *C. freundii* strains, CF003 and CF004 (shown as blue), to all *C. freundii* genomes deposited at GenBank and PATRIC were analysed and drawn into two trees, A and B. Genomes belonging to the same sub-clade with the closest evolutionary distance are coloured as red whilst isolates from South Africa are shown as green (A). The various phylogenetic clades and sub-clades are highlighted together and uniquely to show their evolution and epidemiology. The trees were drawn using the maximum-likelihood method in RAxML, using *Streptomyces sp*. MUSC164 as reference.

The *E. hormaechei* strains were clustered into three, with the *mcr-9.1* strains being very closely related within the same sub-clade; K006 and K063 were of the same clone. Moreover, EC001 and EC015 were of the same clone just as EC009 and EC010 (Fig. 4A). However, EC001 and EC015 were distantly related in the presence of more closely related genomes (Fig. 4B-C). The *mcr-9.1-*positive strains were of close evolutionary relationship with other *E. hormaechei* strains from China, UK, USA, and Thailand: **EB_P9_L5_03**.**19** [*aac(6’)-Ib4, aph(3’’)-Ib, aph(6)-Id*, ***bla***_**ACT-56**_, ***bla***_**IMP-1**_, ***bla***_**TEM-1**_, *catA, catA2, dfrA19, fosA* ***mcr-9***.***1***, *qacE, qacEdelta1, qnrA1, sul1*], **2011WA-NCV** [***bla***_**ACT-56**_, *catA, fosA*], **2011WA-SCV** [***bla***_**ACT-56**_, *catA, fosA*], **35415** [***bla***_**ACT-43**_ *catA, fosA, OqxB*], **WCHEH090020** [***bla***_**ACT-56**_, *catA, fosA*], **UBA4405, WCHEH090006** [***bla***_**ACT-56**_, *catA, fosA*], **T38-C141** [*aph(3’’)-Ib, aph(6)-Id*, ***bla***_**ACT-17**_, ***bla***_**CMY**_, ***bla***_**OXA-48**_, *bleO, catA, dfrA14, fosA*, ***mcr-1***.***1, mcr-3***.***1***, *sul2, tet*(A)], **UBA6755**, S6 [*aac(6’)-Il, aadA1, aadA2, aph(3’’)-Ib, aph(6)-Id*, ***bla***_**ACT-27**_, ***bla***_**SHV-12**_, ***bla***_**TEM-1**_, ***bla***_**VIM-1**_, *catA, dfrA1, dfrA12, fosA*, ***mcr-9***.***1***, *mph(A), oqxB, qacE, qacEdelta1, qnrA1, sul1*], and **UBA5648** (Fig. 4A-B).

**Figure 4.**
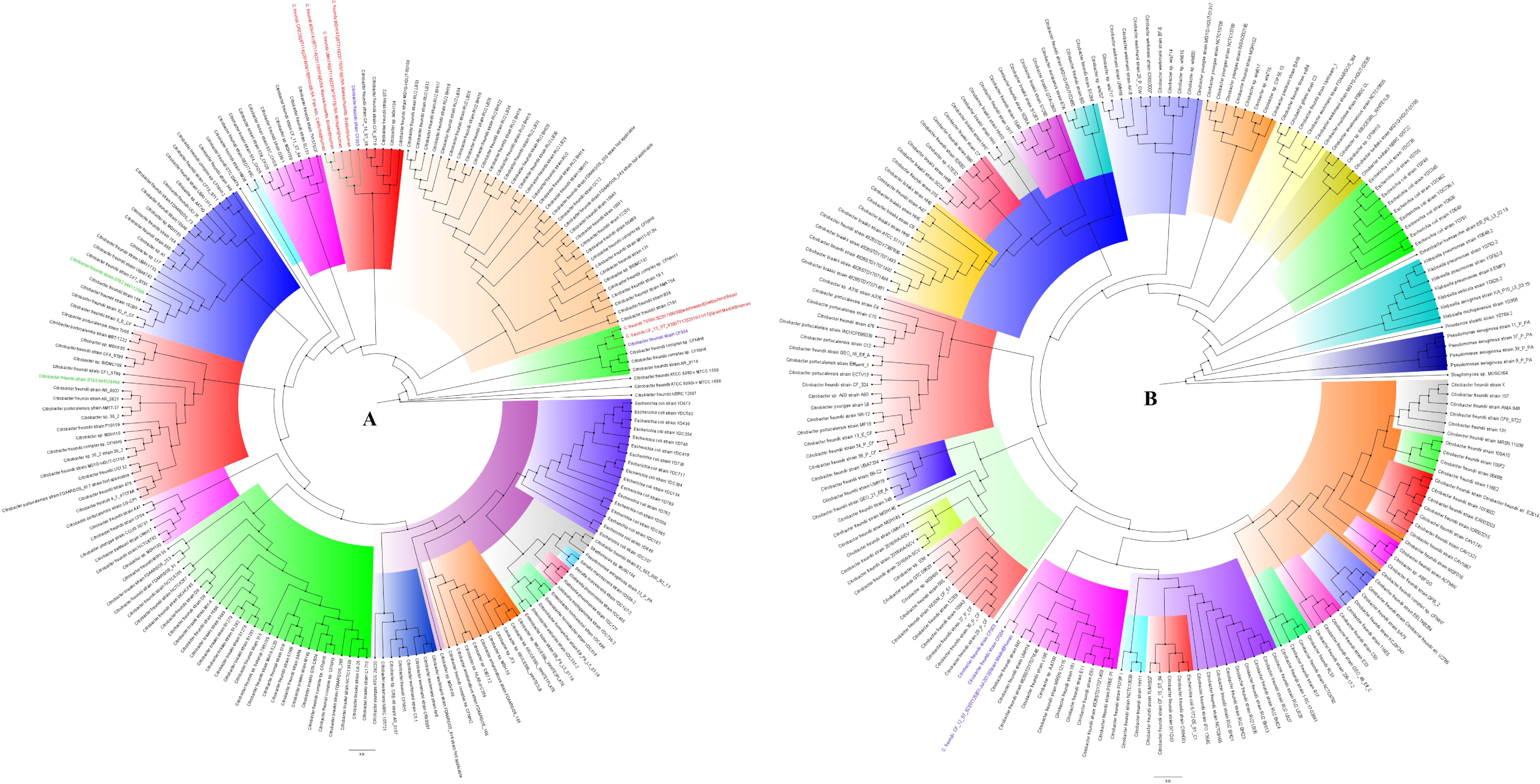
Global phylogenomics of *Enterobacter hormaechei* strains obtained from PATRIC/GENBANK. The relationship of the seven *E. hormaechei* strains, EC001, EC009, EC010, EC015, K006, K063 and K130 (shown as blue), to all *E. hormaechei* genomes deposited at GenBank and PATRIC were analysed and drawn into three trees, A, B and C. Genomes belonging to the same sub-clade with the closest evolutionary distance are coloured as red. The various phylogenetic clades and sub-clades are highlighted together and uniquely to show their evolution and epidemiology. The trees were drawn using the maximum-likelihood method in RAxML, using *Streptomyces sp*. MUSC164 as reference.

The closely related EC009 and EC010 strains were closely related to Chinese and American strains i.e., **WCHEH085055** [*aac(3)-IId, aadA5*, ***bla***_**ACT-41**_, ***bla***_**IMP-4**_, ***bla***_**TEM-1**_, *catA, catA2, dfrA17, fosA, mph(A), oqxB, qacE*Δ*1, qnrS1, sul1, tet*(D)], **UBA7898** and **GN04900** [***bla***_**ACT- 41**_ *catA, fosA, OqxB*]. EC001 and EC015 were of a close phyletic group with strains from USA, China, Croatia, Spain, and Turkey. For EC001, **UBA6646, SMART_488** [*aac(6’)-Ib-cr5, aph(3’’)-Ib, aph(3’)-Ia, aph(3’)-VIa, aph(6)-Id*, ***bla***_**ACT**_, ***bla***_**CTX-M-15**_, ***bla***_**KPC-2**_, ***bla***_**OXA-1**_, ***bla***_**TEM-1**_, *catA, catA2, catB3, dfrA14, fosA, oqxB, qnrB19, sul2, tet*(A)], **SMART_723** [*aac(3)-II, aac(6’)-IIc, aph(3’’)-Ib, aph(6)-Id, arr*, ***bla***_**ACT-45**_, ***bla***_**DHA-7**,_ ***bla***_**OXA-48**_, ***bla***_**SHV-12**_, ***bla***_**TEM-1**_, *catA, catA2, dfrA19, ere(A), fosA*, ***mcr-9***.***1***, *oqxB, qacE, qnrB4, sul1*] and **WCHEX090075** [*aadA2, ant(2’’)-Ia*, ***bla***_**ACT**_, ***bla***_**NDM-1**_, ***bla***_**SHV-12**_, *ble, catA, catA1, dfrA14, fosA*, ***mcr-9***, *oqxB, qacE, qnrA1, qnrS1, sul1*] were of the same clade whilst **WCHEH090043** [***bla***_**ACT**_, *catA, fosA, oqxB*], **UBA1647, SMART_530** [***bla***_**ACT**_, ***bla***_**OXA-48**_, *catA, fosA oqxB, qnrS1*], **SMART_1112** [*aac(6’)-Ib4, aac(6’)-Im, aadA1, aadA16, aph(2’’)-IIa, aph(3’’)-Ib, aph(3’)-Ia, aph(6)-Id, arr-3*, ***bla***_**ACT**_, ***bla***_**CTX-M-15**_, ***bla***_**TEM-1**_, ***bla***_**VIM-1**_, *catA, catA2, dfrA19, dfrA27, fosA*, ***mcr-9***.***1***, *oqxB, qacE*Δ*1, qnrB6, sul1, sul2, tet(A), tet*(D)], **SMART_562** [*aac(6’)-Ib4, aac(6’)-Im, aadA1, aadA16, aph(2’’)-IIa, aph(3’’)-Ib, aph(3’)-Ia, aph(6)-Id, arr-3*, ***bla***_**ACT**_, ***bla***_**CTX-M-15**_, ***bla***_**TEM-1**_, ***bla***_**VIM-1**_, *catA, catA2, dfrA19, dfrA27, fosA*, ***mcr-9***.***1***, *oqxB, qacE*Δ*1, qnrB6, sul1, sul2, tet(A), tet*(D)] etc. clustered together (Fig. 4).

The closest evolutionary relative of K001 was a single strain, **WUSM_KV_44** [***bla***_**LEN-13**_, *fosA, oqxA, oqxB*] from the USA (Figure 5) whilst PM005, which was initially identified as *Proteus mirabilis* by MicroScan, but later revised by NCBI’s ANI to *P. alcalifaciens*, was not related to any *Providencia sp*. genome (Figure 6A-B); however, it was closely related to *Proteus mirabilis* **Pr2921** strain from Uruguay [*catA, tet(J)*] (Figure 6C).

**Figure 5.**
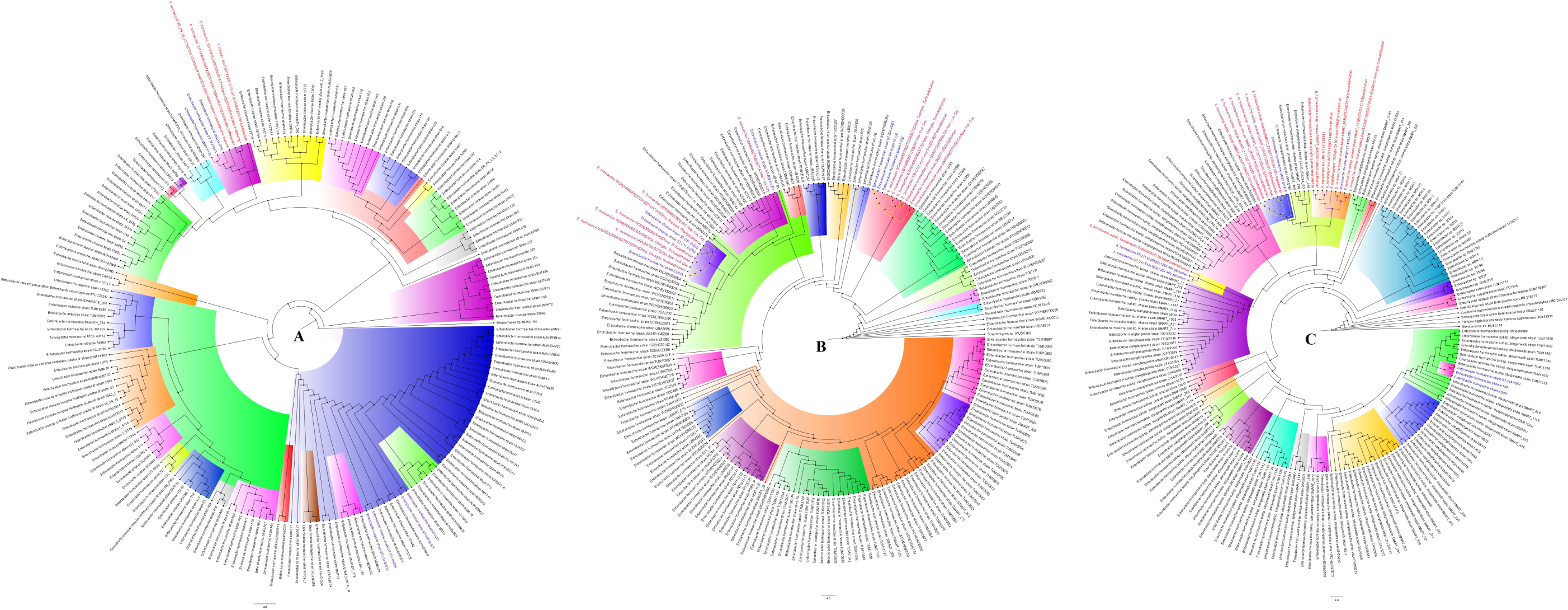
Global phylogenomics of *Klebsiella variicola* strains obtained from PATRIC/GENBANK. The relationship of the *K. variicola* strain, K001 (shown as blue), to all *K. variicola* genomes deposited at GenBank and PATRIC were analysed and drawn into two trees, A and B. Genomes belonging to the same sub-clade with the closest evolutionary distance are coloured as red. The various phylogenetic clades and sub-clades are highlighted together and uniquely to show their evolution and epidemiology. The trees were drawn using the maximum-likelihood method in RAxML, using *Streptomyces sp*. MUSC164 as reference.

**Figure 6.**
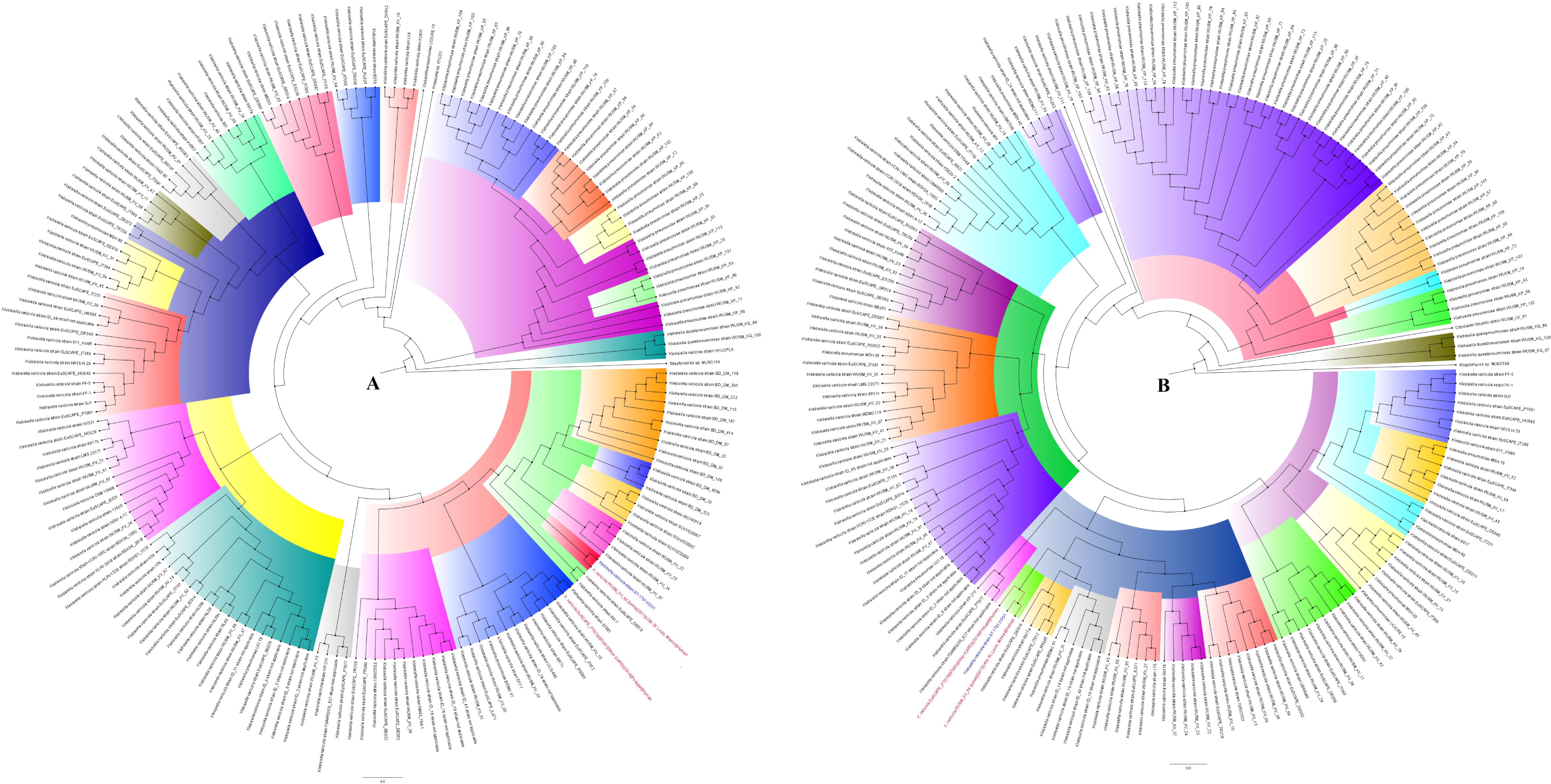
Global phylogenomics of *Providencia alcalifaciens* and *Proteus mirabilis* strains obtained from PATRIC/GENBANK. The relationship of the *P. alcalifaciens* strain, PM005 (shown as blue), to all *Providencia* (A and B) *and Proteus* (C) genomes deposited at GenBank and PATRIC were analysed and drawn into two trees, A, B and C. Genomes belonging to the same sub-clade with the closest evolutionary distance are coloured as red. The various phylogenetic clades and sub-clades are highlighted together and uniquely to show their evolution and epidemiology. The trees were drawn using the maximum-likelihood method in RAxML, using *Streptomyces sp*. MUSC164 as reference.

## Discussion

We herein report on the first emergence of *mcr-9.1* in both South Africa and Africa, in three *E. hormaechei* strains with a rich repertoire of resistomes and mobilomes. The *mcr-9.1* strains were collected among clinical Enterobacteriaceae strains during a molecular surveillance procedure to identify ESBL producers in a referral laboratory. It is a worrying observation to have all the strains, sourced from different patients and wards within the same hospital, being multi-drug resistant. Although the resistomes largely reflected the resistance phenotype, resistance to antibiotics were observed without an underlying resistance gene being identified. Specifically, the *C. freundii* strains expressed resistance to several antibiotics although not more than three resistance genes were found in them. This could suggest the presence of an unknown resistance determinant or the use of active efflux. Furthermore, sensitivity to certain antibiotics such as colistin was not corroborated by the presence of *mcr-9.1*. Sensitivity to colistin in strains having the *mcr* gene has been reported previously and chromosomal mutations have been shown to exert a higher resistance MIC than *mcr* genes ^3,25–27^.

Only a single strain with the *mcr* gene was colistin resistant and the identified mutations in *mgrB* and *pmrAB* were also found in susceptible strains. Hence, the identified mutations cannot be responsible for colistin resistance. Consequently, other factors identified for colistin resistance or other unknown determinants could be responsible for the observed colistin resistance ^3,7,11,28^. The three strains bearing the *mcr-9.1* gene were closely related, but the contigs bearing the *mcr-9.1* genes were not of the same sequence homology (Supplemental data 2), with the *mcr-9.1-*contigs’ distance trees showing that K006 and K130 were closer (Fig. 2B). It is worth noting that K006 and K130 were phylogenetically distant, but their *mcr-9.1-*contigs’ were of closer nucleotide identity than K006 and K063, which were of the same clone (Fig. 4). This observation, plus the close alignment of the *mcr-9.1-* contigs’ with plasmid genomes on Genbank (Fig. 2B), strongly suggest that the *mcr-9.1* gene could have been horizontally, instead of vertically, acquired.

The global evolutionary trajectory of the *mcr-9* gene and genomes shows six major phylogenetic clades, herein labelled A to F. The strains and plasmids in clade A seem to be the earliest ancestors of this gene with F being the last and most recent. Moreover, *Enterobacter spp*., particularly *E. hormaechei*, and *Salmonella enterica* plasmids are the commonest hosts of the *mcr-9* gene (Fig. 2B). This is not surprising as *mcr-9.1* was first identified in most *Salmonella* Typhimurium ^25^. The diversity of species and plasmids involved in the dissemination of this gene explains its promiscuity and rapid spread around the globe, further corroborating the need to restrict the use of colistin in both veterinary and human medicine ^5,29^.

The genetic support of the various resistance genes, particularly *bla*_CTX-M-15_, *bla*_TEM-1_, *bla*_OXA_ and *mcr-9.1* identified in these isolates are not new. In particular, the co-occurrence of *bla*_*CTX- M-15*_ and *bla*_*TEM-1*_ within Tn*3* composite transposons, IS*Ec9* and IS*19* on IncF type plasmids is widely reported in both South Africa and worldwide ^4,20–22,30,31^. Further, the presence of the *mcr-9.1* gene on IncHI2 plasmids as well as the presence of a cupin fold metalloprotein, as observed here, are common around *mcr-9.1* genes ^25^. The promiscuity of IncF plasmids and the abundance of integrons, transposons and ISs in the genomes of these strains is worrying as they might mobilise and facilitate the faster dissemination of these multiple resistance genes to other species and clones ^4,31,32^.

It is interesting to note that not all the closely related strains around the *mcr-9.1* strains harboured the *mcr* gene. Some of the global strains with very close phyletic clustering with the non-*mcr-*positive strains contained *mcr*, and in some cases, carbapenemase (*bla*_NDM_, *bla*_OXA-48_, *bla*_VIM_ and *bla*_IMP_) genes. Such resistome differences between strains belonging to the same clade suggest the gain and loss of resistance plasmids during the evolutionary and epidemiological trajectory. This could be the case for the other *E. hormaechei* strains that had no *mcr* gene. The 11 strains included in this study were phylogenetically distant from any strain from South Africa or Africa, but of very close evolutionary distance to international strains, some of which harboured very rich resistomes, including the co-occurrence of carbapenemases and *mcr* genes. Thus, the possibility of these strains having been imported cannot be ruled out, a situation that warrants constant surveillance and screening of medical tourists, particularly those from carbapenemase- and *mcr-*endemic areas ^6,33^.

Although carbapenemases were not found in these strains as have been reported elsewhere in *mcr-*positive strains ^34–38^, the co-occurrence of *mcr* and ESBL genes is worrying as they restrict therapeutic options. To date, only *mcr-1* genes have been reported in South Africa ^6,27,39,40^, making the emergence of *mcr-9.1* in strains as old as 2013 a worrying situation. This suggests that other *mcr* variants could be present in South Africa and Africa, and intensive clinical surveillance is necessary to unearth these and pre-empt further escalation.

## Data Availability

All data on this manuscript are available as supplemental information attached to this article.

## Acknowledgement

none

## Transparency declaration

authors declare no conflict of interest

## Author contributions

JOS designed and supervised the study, undertook all the analyses, bioinformatics, image designs and manuscript write-up; NEM, LM and NMM undertook the laboratory assays; all authors proof-read and agreed to the final version of the manuscript.

## Ethical considerations

All protocols and consent forms were executed according to the agreed ethical approval terms and conditions. All clinical samples were obtained from a reference laboratory and not directly from patients, who agreed to our using their specimens for this research. The guidelines stated by the Declaration of Helsinki for involving human participants were followed in the study.

---

**Figure S1.**
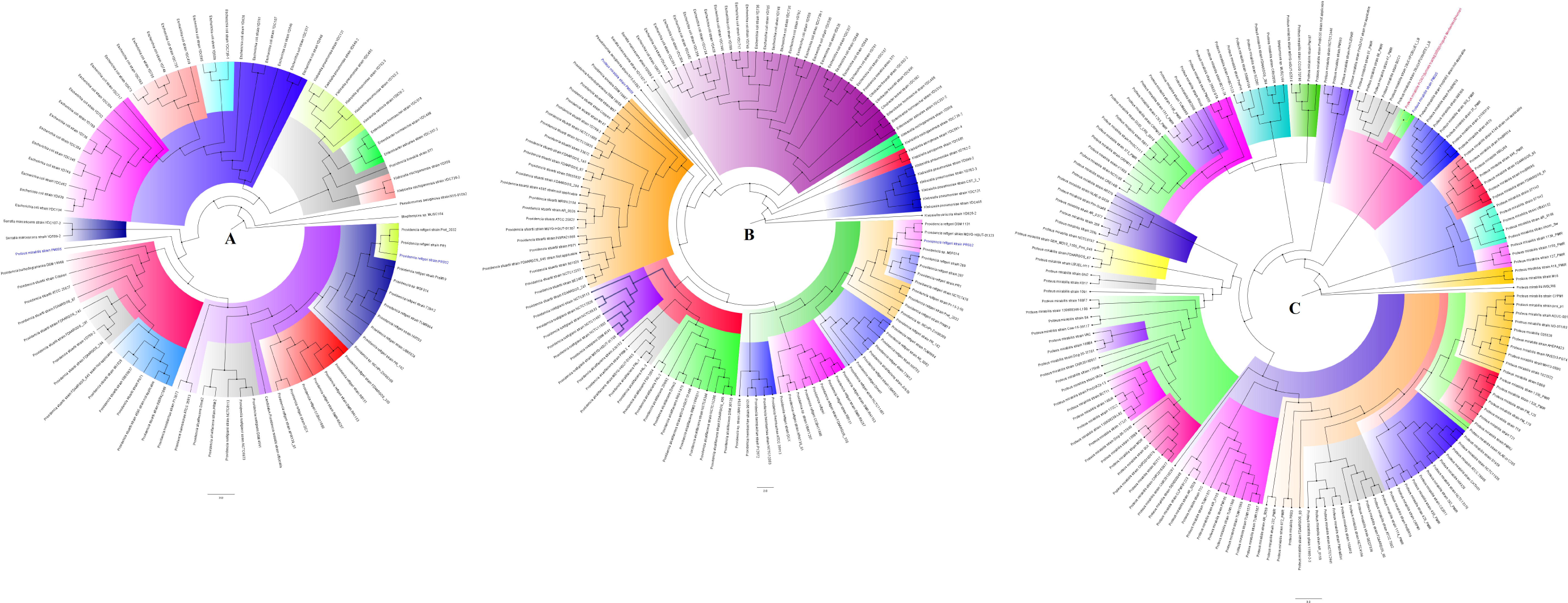
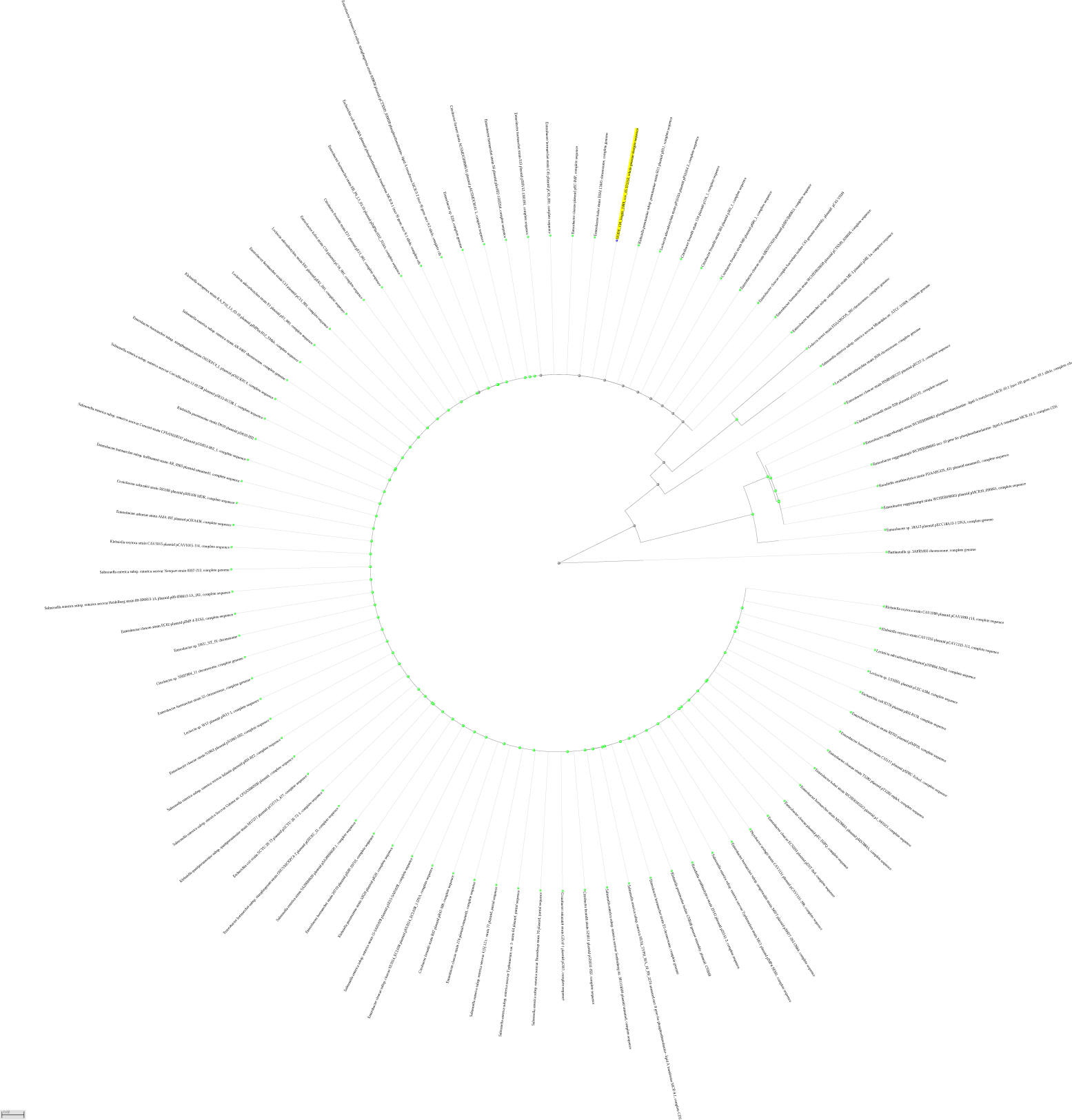
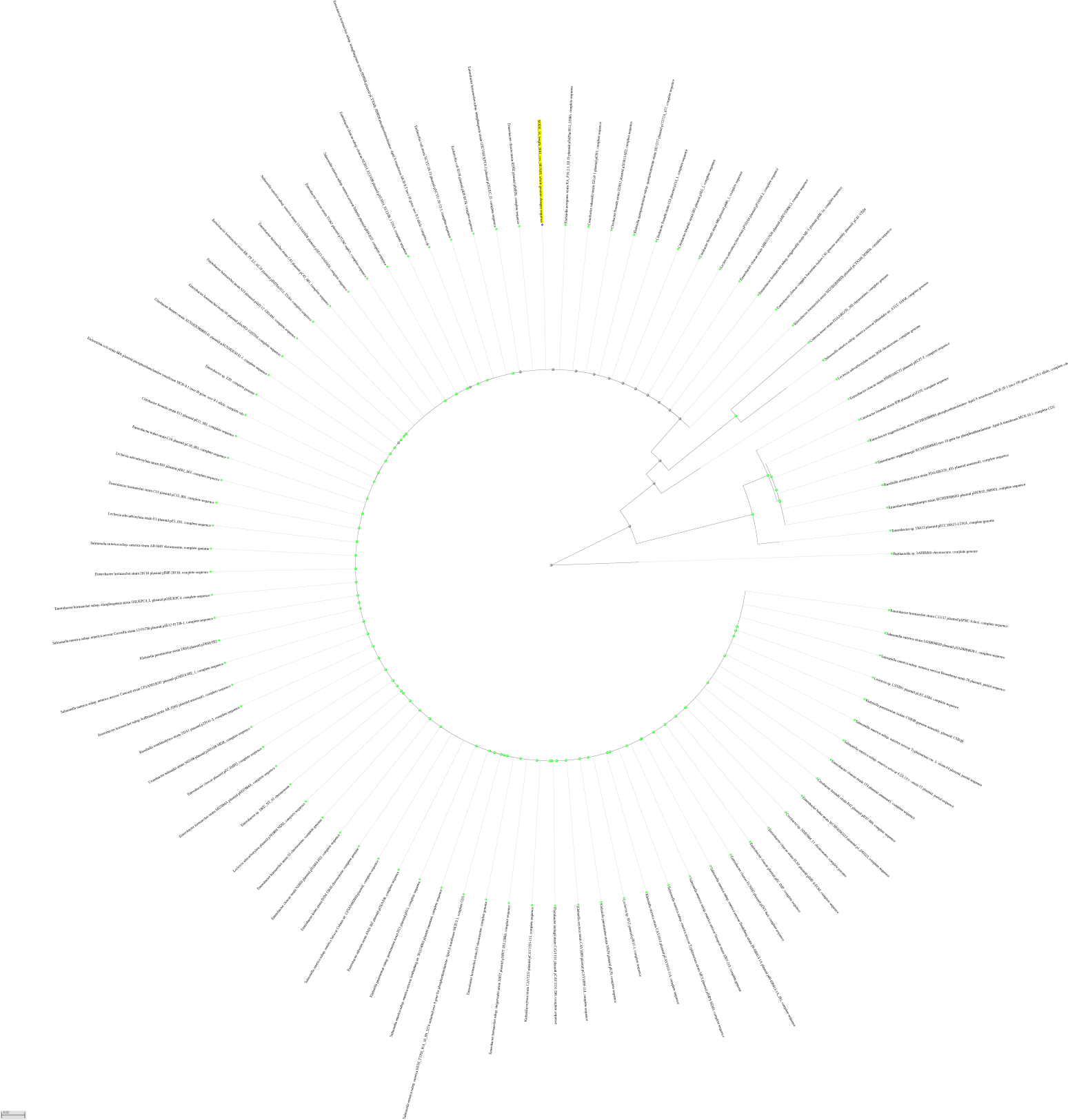
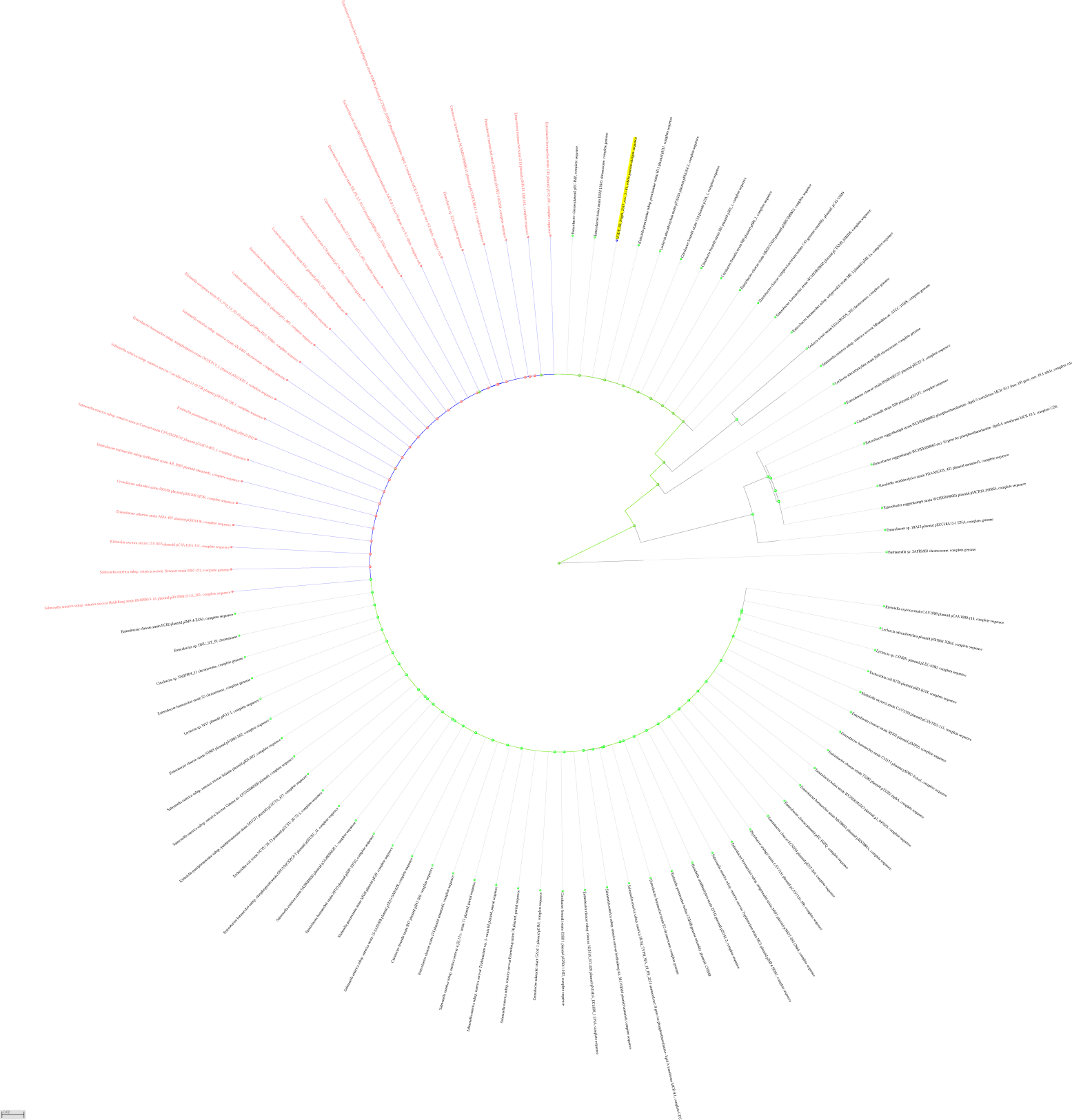
The evolutionary epidemiology of *mcr-9.1* genes contained on different plasmids and genomes worldwide. A PDF version of Figure 2A that can be zoomed to clearly see the plasmids and genomes bearing the *mcr-9.1* genes are shown for K006 (A), K063 (B) and K130 (C), which are coloured or highlighted as yellow.

**Supplemental Table S1**. Antimicrobial sensitivity results and resistome of the isolates. The sensitivity of the isolates to the various antibiotics tested using the MicroScan is shown, with those coloured as green being resistant according to the CLSI breakpoints. Those coloured as blue are resistant according to the EUCAST breakpoints. Those not coloured are susceptible. The various antibiotic classes to which the antibiotic agents belong are shown above each antibiotic in unique colours and the resistance genes per isolate are shown in the last column.

**Supplemental dataset 1**. General data of demographic, phenotypic and genomic results used for this study.

**Supplemental dataset 2**. Sequences and alignment of contigs harbouring the *mcr-9.1* genes.

